# Analysis of the time and age dependence of the case-fatality-ratio for COVID-19 in seven countries with a high total-to-positive test ratio suggests that the true CFR may be significantly underestimated for the United States in current models

**DOI:** 10.1101/2020.05.13.20101022

**Authors:** Jessica Rothman, David Eidelberg, Samantha Rothman, Theodore Holford, Douglas L Rothman

## Abstract

**Background:** Knowing the true infected and symptomatic case fatality ratios (IFR and CFR) for COVID-19 is of high importance for epidemiological model projections. A large correction factor is usually applied for missed cases. For the United State reported CFR of 5.96%, the estimated IFR values are 10–50-fold lower, justified by early reported CFR values of 0.1% to 0.5% in countries with more extensive testing. However, since then these values have risen 5 to 10-fold. We analyzed their age dependent CFR time courses to explain this increase and to determine whether a common factor can explain their CFRs.

**Methods:** Age dependent time to fatality corrected CFR was calculated using two independent methods. A linear model was developed that predicts CFR based on age dependent CFR coefficients and the age distribution of cases. The model was tested by a linear regression of each country’s CFR against case percentage of 70 years and over. The model was further tested by calculating the percent of the population in New York City who have been infected.

**Results:** Corrected CFR values ranged from 0.58% to 5.0%. The large majority of CFR variation was explained by case age distribution above 70 years old. Using the CFR derived from the linear model we predicted between 14.7% and 22% of the adult population in NYC had been infected by COVID-19, in agreement with random testing studies (15.3% – 21%).

**Conclusions:** The large rise in the reported CFR is due to the delay time between infection/diagnosis and fatality with COVID-19. The linear model based on their age specific CFR values provides an alternative method for calculating the true CFR in other regions. Most of the variation in CFR between countries was dependent on case age distribution, which must be considered in measures for mitigating the extensive impacts of the pandemic.

## Introduction

Knowing the fraction of individuals infected with COVID-19 who will likely die or require hospitalization is critical for epidemiological modeling and public health policy. Obtaining an accurate estimate of the fatality ratio for all symptomatic cases (referred to as the case fatality ratio, CFR) and infected cases (IFR) is complicated by mild and asymptomatic cases not being detected, and by the time lag between diagnosis and fatality.(1) For COVID-19, the World Health Organization (WHO) reported a non-corrected CFR (often referred to naïve CFR or nCFR) of 3.5 +/− 0.2 % as of March 3, 2020.(2) Several reports have attempted to estimate the true IFR and CFR from this data as well as from the data of other countries. As shown in Table 1, with one exception, the true CFR values for China were estimated to be between 0.85% and 1.4% which are all substantially lower than the reported Chinese nCFR value.(3–7) For the United States and United Kingdom, even lower true CFR and IFR values have been reported ranging from 0.125% to 0.9%.(3,5,8–11) Support for these lower values has been provided by comparison with reported nCFR values of less than 1% early in the outbreak from countries that were believed to have captured almost all of their symptomatic cases due to extensive testing and tracing of contacts.(4,8,9,12) Another cited example is the Diamond Princess cruise ship, in which all passengers were tested, for which early nCFR/IFR reports were in the 0.3% to 1.1% range.(4,6,8,9)

**Table 1:**
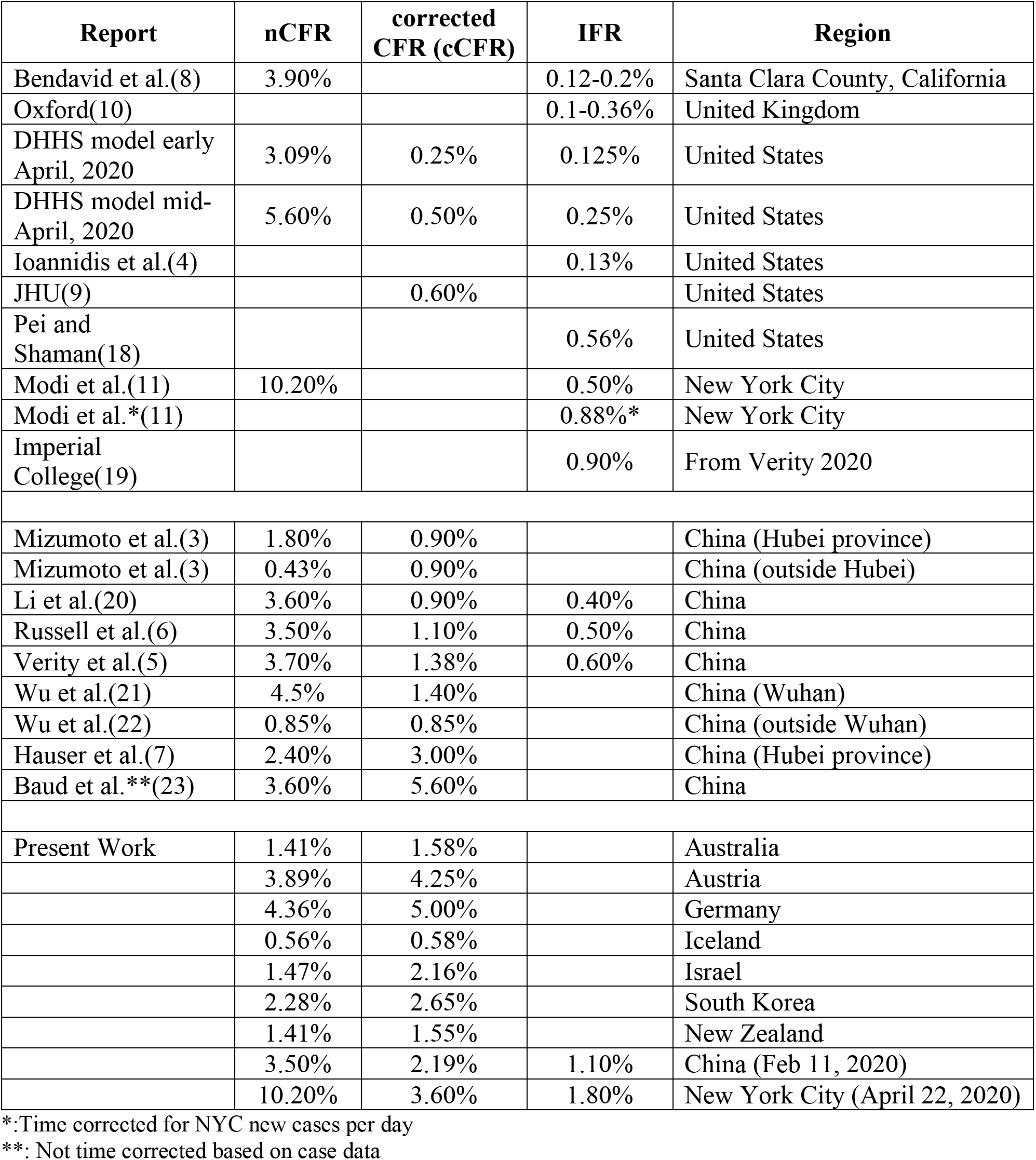
Reported nCFR, corrected CFR, and corrected IFR values for China, the United Kingdom and the United States. The table summarizes reported corrected CFR and IFR values, the nCFR value at the time of the report, and the country/region. Details are available in the cited references. (3–11,13,20–29) Studies are listed by their first author or by the location of the modeling group that reported them. Abbreviations: DHHS: Department of Health and Human Services, USA; Oxford: Oxford College, U.K.; Imperial College: Imperial College, U.K.

| Report | nCFR | corrected CFR (cCFR) | IFR | Region |
| --- | --- | --- | --- | --- |
| Bendavid et al.(8) | 3.90% |  | 0.12-0.2% | Santa Clara County, California |
| Oxford(10) |  |  | 0.1-0.36% | United Kingdom |
| DHHS model early April, 2020 | 3.09% | 0.25% | 0.125% | United States |
| DHHS model mid-April, 2020 | 5.60% | 0.50% | 0.25% | United States |
| Ioannidis et al.(4) |  |  | 0.13% | United States |
| JHU(9) |  | 0.60% |  | United States |
| Pei and Shaman(18) |  |  | 0.56% | United States |
| Modi et al.(11) | 10.20% |  | 0.50% | New York City |
| Modi et al.*(11) |  |  | 0.88%* | New York City |
| Imperial College(19) |  |  | 0.90% | From Verity 2020 |
| Mizumoto et al.(3) | 1.80% | 0.90% |  | China (Hubei province) |
| Mizumoto et al.(3) | 0.43% | 0.90% |  | China (outside Hubei) |
| Li et al.(20) | 3.60% | 0.90% | 0.40% | China |
| Russell et al.(6) | 3.50% | 1.10% | 0.50% | China |
| Verity et al.(5) | 3.70% | 1.38% | 0.60% | China |
| Wu et al.(21) | 4.5% | 1.40% |  | China (Wuhan) |
| Wu et al.(22) | 0.85% | 0.85% |  | China (outside Wuhan) |
| Hauser et al.(7) | 2.40% | 3.00% |  | China (Hubei province) |
| Baud et al.** (23) | 3.60% | 5.60% |  | China |
| Present Work | 1.41% | 1.58% |  | Australia |
|  | 3.89% | 4.25% |  | Austria |
|  | 4.36% | 5.00% |  | Germany |
|  | 0.56% | 0.58% |  | Iceland |
|  | 1.47% | 2.16% |  | Israel |
|  | 2.28% | 2.65% |  | South Korea |
|  | 1.41% | 1.55% |  | New Zealand |
|  | 3.50% | 2.19% | 1.10% | China (Feb 11, 2020) |
|  | 10.20% | 3.60% | 1.80% | New York City (April 22, 2020) |

\*:Time corrected for NYC new cases per day
\*\*: Not time corrected based on case data

To determine whether these countries continued to have a low nCFR later in their outbreaks, we reexamined the reported number of fatalities and nCFR values outbreaks on May 7, 2020 for Australia, Austria, Germany, Iceland, Israel, New Zealand, and South Korea. In all cases these countries continued to have a high degree of testing and tracking and testing of contacts as shown by their ratio of total tests to positive tests ranging from 16:1 to 127:1, versus 7:1 for the United States and 8:1 for the United Kingdom, (Appendix 4). Therefore, their final reported nCFR values, when all cases are complete, are potentially close to their true CFR value (if all symptomatic cases were tested). To determine why there was a large increase in their reported CFR over time we calculated for each country a CFR values corrected for the known time delay between diagnosis (time of test) and fatality (corrected CFR) using two methods.(3) In the standard approach, we used a probability distribution function (*f_D_*) for the estimated probability of a fatality versus “days-after-diagnosis”.(1,3,6) We also independently estimated the corrected CFR from the closed case CFR time course. The methods gave similar corrected CFR values for each country (Table 2). Furthermore, our simulations showed that both methods provide accurate CFR values even early in the outbreaks when the reported nCFR values were several fold lower.

**Table 2.** Comparison of the corrected CFR values calculated using the closed case CFR and time to fatality correction methods. (12–14,24–27,29)

| Country | Closed Case CFR | Time- Corrected CFR |
| --- | --- | --- |
| Australia | 1.58 | 1.42 |
| Austria | 4.26 | 4.20 |
| Germany | 5.02 | 5.05 |
| Iceland | 0.57 | 0.58 |
| Israel | 2.16 | 1.72 |
| New Zealand | 1.55 | 1.51 |
| South Korea | 2.65 | 2.32 |

As shown in Table 1, for the countries examined, despite early low reported nCFR values and very high levels of testing (Appendix 4) we calculated a wide range of corrected CFR values – from 0.6% to 5.0%. These values are well above the majority of CFR and IFR values estimated for the United States and United Kingdom.(5,10,13,14) Despite this range, the large majority of the variation between countries was explained by the fraction of each country’s cases in the age ranges of 70 – 79 and 80 and above (Table 1, Figure 6). These age groups accounted on average for over 80% of the reported fatalities. The finding of a strong correlation of the corrected CFR with age also supports the hypothesis that the testing in each country was sufficient to capture the majority of their symptomatic cases.

Using the corrected CFR for these groups and the population under 70, we developed a linear model for predicting the CFR and IFR values for countries and regions with limited testing. From this model we estimated the corrected CFR for China using the linear model from the case age distribution data reported February 11, 2020 (WHO release) at 2.19% (95% CI of the mean: 1.54%-2.85%). As shown in Table 1, this value is higher than the majority of previous estimates for China, but falls within their range when earlier values are multiplied by the 1.5 fold increase in total number of fatalities recently reported by the Chinese government.(2)

Regarding US and UK populations, there is a much wider range of CFR and IFR estimates, with the lowest values being almost 40 times below the highest CFR we calculated (Germany).(4,5,8–11) We therefore tested our model by calculating the percent of the population who has been infected with COVID-19 in New York City and comparing it to the results of recent random testing studies. New York City was chosen because it is undergoing a large outbreak, and it is likely, therefore, that the percentage of infected individuals is sufficiently high such that false positives and negatives in antibody tests used would not have a large effect on the study’s outcome.(15,16) We predicted as of April 22, 2020 the percentage of the adult population that had been infected by COVID- was between 4.69% and 22.01%. These values are consistent with the reported values of 15.3% and 21%.(16,17) In contrast, using previously reported IFR values gave minimum estimates between 1.5 and 10-fold higher (see Figure 7).(5,8–11,15,16)

## Sources of data

Data were obtained from data compiling sites (Worldometer, Statista) and the Australian, Austrian, German, Iceland, Israeli, South Korean, and New Zealand government websites.(12–14,24–27,29,30) Data was also obtained from the New York City Department of Health website.(17,31)

## Definitions

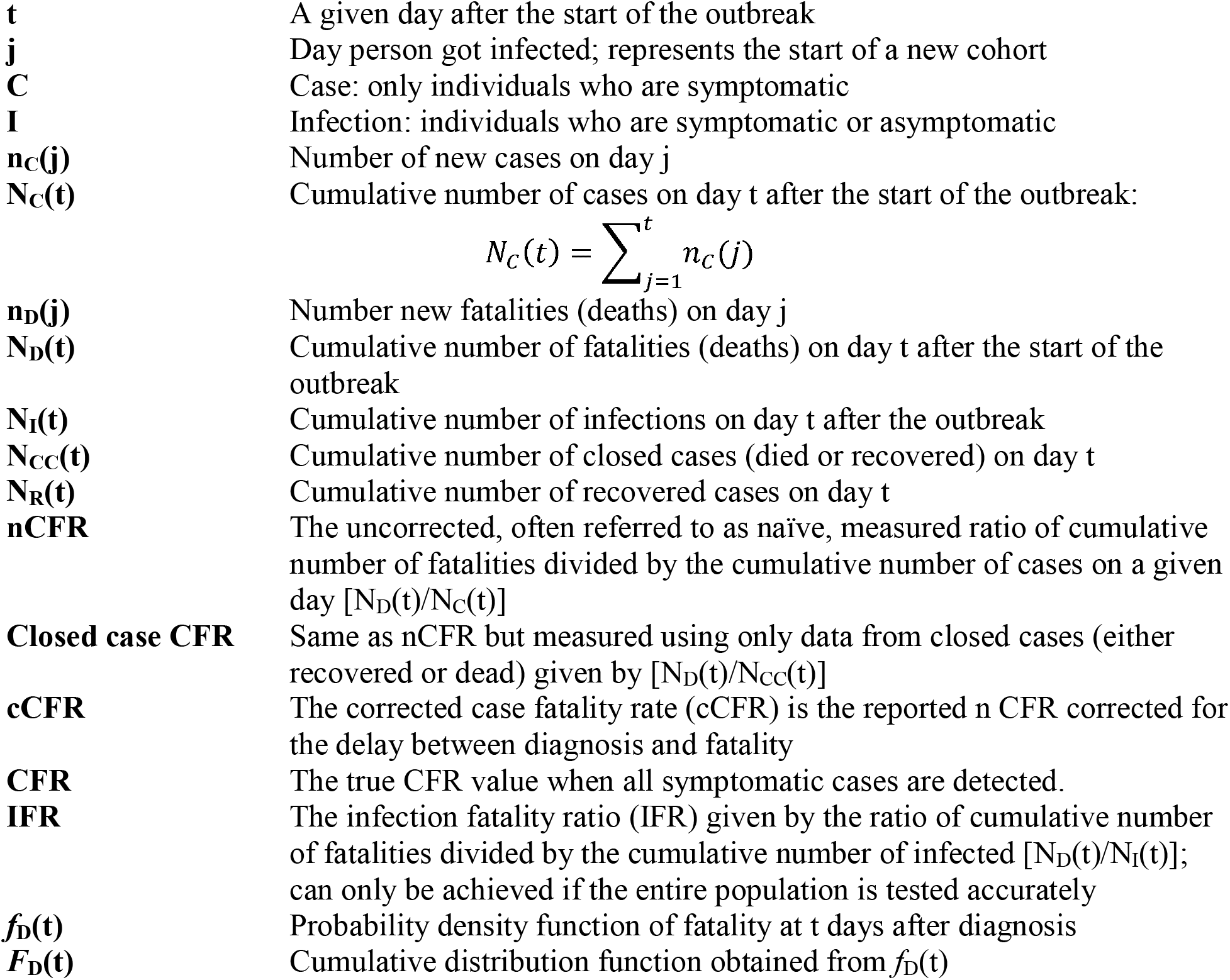

## Calculations

### Estimation of the corrected CFR based on correction of the reported nCFR(t) for the delay between diagnosis and fatality and from the closed case CFR

We used two independent methods to estimate the corrected CFR. In one method we corrected the reported nCFR(t) for the time delay between diagnosis and fatality based on previously reported approaches.(3,5–7,21,23,32,33) The second method was based on our observation that in all countries analyzed the closed case CFR (see definitions) converged to a near constant value prior to the nCFR. A closed case is defined as a case that has been designated as recovered or has died. Unlike the nCFR, it is not impacted by changes in the percentage of the infected population that is tested during the duration of the epidemic. As shown in Appendix 3, provided that the median times to fatality and for recovery stay approximately constant during the outbreak, the closed case CFR will converge to the final value prior to the nCFR.

We implemented a time-delay-to-fatality correction method using an f_D_ derived from reported log-normal fits of data obtained from China, between December and late January, of the percentage of fatalities of COVID-19 patients per day after diagnosis.(1,3,5,6,33) Data was used only from patients who were hospitalized outside of Hubei province to avoid the potential problem that adequate medical care was likely not available within the province, and especially in Wuhan, early in the outbreak.(3,7,21) For the cohort of cases diagnosed on day j, the *f_D_* at day t is described by,

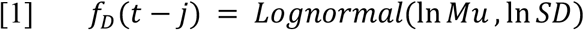

The calculated cumulative number of fatalities from the cohort diagnosed on day j on day t was calculated from the cumulative distribution (*F*_D_) which is the integral of Eq. [1] from day j to day t multiplied by the number of new cases on day j and the corrected CFR,

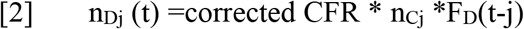

where *t* > *j*.

We note that Eq. [2] is equivalent to a convolution integral of *f_D_*(t-j) with a delta function centered at day j with an area of CFR*n_Cj_.

Total calculated number of fatalities on day t was then obtained by adding together the fatalities from each cohort,

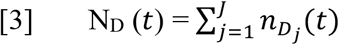

Where J is the total number of cohorts.

We then calculated the predicted value of nCFR(t) on day t using Eq. [3] and the cumulative cases up to that day where N_D_(t) is calculated,

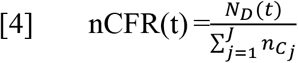

The value of the corrected CFR was then calculated by adjusting the value of the corrected CFR in Eq. [2] until the calculated nCFR(t) on the last day of the outbreak analyzed was equal to the reported value.

### Optimization of the parameters of *f*_D._

The function f_D_ was based on reports of the measured onset (day of positive test) to fatality distributions for Chinese patients outside of Wuhan who were infected in December and January by Linton et al. and Mizumoto et al.(3,33). These investigators modeled the distributions as Log-normal functions that were corrected for right censoring (fatalities missed due to the limited patient observation time). The best fitting distributions from these sources were very similar, with Linton reporting a best fit median of 13.2 days with a 95% CI of 11.5 to 15.3 days, and Mizumoto et al. reporting a best fit median (estimated from their reported log-mean value) of approximately 13 days.(33–35) Because two other studies fitting to a different distribution function reported higher mean and extrapolated median values, we decided to examine median values of 14, 17, and 21 days. The SD reported by all studies, based on gamma fits, was very similar, and equivalent to a logSD of approximately 0.50 as reported by Mizumoto.(34,35)

For the least square fits, we first determined the corrected CFR from the data from Germany using the time correction method described above for each of the distribution functions.

Germany was chosen because of the large number of cases compared to the other countries analyzed; this minimizes statistical fluctuations in the early outbreak of the reported nCFR(t) curve. Goodness of fit was then determined by calculating the least squares total residual by squaring the differences between our calculated nCFR(t) (using the corrected CFR) and the reported nCFR(t) values, and then summing those squares. The optimum parameter values were determined based upon evaluating medians of 14, 17, and 21 days and for each value of the median varying logSD from 0.25 to 0.75.

### Model for calculating the corrected CFR based on the age distribution of positive cases in the population and the age specific CFR values

Studies have reported that the nCFR for COVID-19 strongly increases with age.(5–7,9–11,15,21,22,32–34) We determined for each country the corrected CFR values for the fraction of a country’s populations of age 0–69, 70–79, and 80 and above based on reports of the number of fatalities and number of cases in each age range. The population below 70 years old was not further subdivided due to the relatively low number of fatalities in this group in the countries with the least number of cases. The total reported corrected CFR is then expressed by,

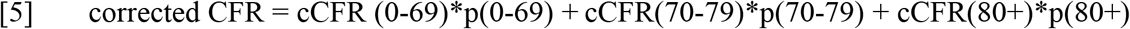

where **p()** is the proportion of the population in the relevant age group and the terms, **cCFR**(), are the corresponding corrected CFR for the population in that age-group. Table 3 gives the values we calculated from the reported data of each country for cCFR(0–69), cCFR(70–79), and cCFR (80+). Under the assumption that the age specific corrected CFR values for each country analyzed was close to the true value, we used Eq. [5] to calculate the true corrected CFR for New York City and China. To perform the calculation, we used the mean coefficients of all countries analyzed for cCFR(0–60), cCFR(70–79), and cCFR(80+), and the reported values. The corrected CFR for New York City and China were then determined based on their reported values of p(0– 69), p(70–79), and p(80+). The IFR was calculated from the corrected CFR values based on the assumption that the true CFR was achieved in the countries analyzed. A factor of 2 was then used to convert the CFR to IFR based on reports of ½ of all COVID-19 cases being asymptomatic and may have escaped detection.(2,31,36,37)

**Table 3:** Age specific fractions of cases, age specific corrected CFR, and contributions of each age group to the overall corrected CFR for each country. Definitions: **p()** is the proportion of the population in the relevant age group; **cCFR(80+)** is the cCFR for cases 80 years old and above; **cCFR(70–79)** is the cCFR for cases 70 – 79 years old; **cCFR(70+)** is the cCFR for cases 70 years old and above; **cCFR(0–69)** is the cCFR for all cases 69 years old and below; **cCFR70+** is the contribution to the overall cCFR from all cases 70 years old and above; **cCFR0–69** is the contribution to cCFR from all cases 69 years old and below. The **subscript A** refers to cCFR70+ values corrected to have a fraction of 40% of cases 80 years old and above. The value was chosen to match the mean from all countries except New Zealand (which has not reported this value and therefore it was assumed to be the same as the mean of the other countries). Data was obtained from the following references.(1–8)

| Country | p(0-69) | p(70-79) | p(70+) | p(80+) | p(80+)/p(70+) | cCFR(80+) | cCFR(70-79) | cCFR(70+) | cCFR(0-69) |
| --- | --- | --- | --- | --- | --- | --- | --- | --- | --- |
| Australia | 86.11% | 10.54% | 13.89% | 3.35% | 24.09% | 27.50% | 3.95% | 9.62% | 0.28% |
| Austria | 81.51% | 8.62% | 18.49% | 9.87% | 53.37% | 24.23% | 12.62% | 18.80% | 0.95% |
| Germany | 81.12% | 8.87% | 18.88% | 10.01% | 53.00% | 31.83% | 13.06% | 23.01% | 0.81% |
| Iceland | 95.38% | 3.40% | 4.62% | 1.22% | 26.51% | 18.95% | 5.13% | 8.79% | 0.18% |
| Israel | 91.00% | 5.30% | 9.00% | 3.70% | 41.11% | 35.30% | 9.49% | 20.10% | 0.39% |
| New Zealand | 92.21% | 7.79% | 7.79% |  |  |  |  | 17.08% | 0.22% |
| South Korea | 88.90% | 6.59% | 11.10% | 4.51% | 40.59% | 28.07% | 12.05% | 18.56% | 0.63% |
| Mean | 88.03% | 7.30% | 11.97% | 5.44% | 39.78% | 27.65% | 9.38% | 16.57% | 0.49% |
| SD | 5.40% | 2.41% | 5.40% | 3.65% | 12.52% | 5.72% | 3.97% | 5.35% | 0.30% |
| 95% CI of Mean |  |  |  |  |  | (21.64%, 33.65%) | (5.22%, 13.55%) | (10.95%, 22.18%) | (0.18%, 0.81%) |

**Table 3: Age specific fractions of cases, age specific corrected CFR, and contributions of each age group to the overall corrected CFR for each country.**
| Country | cCFR<sub>0-69</sub> | cCFR<sub>70+</sub> | cCFR | cCFR<sub>70+</sub>/cCFR | cCFR(70+)<sub>A</sub> | cCFR<sub>0-69A</sub> | cCFR<sub>70+A</sub> | cCFR<sub>A</sub> | cCFR<sub>70+A</sub>/cCFR<sub>A</sub> |
| --- | --- | --- | --- | --- | --- | --- | --- | --- | --- |
| Australia | 0.24% | 1.34% | 1.58% | 84.62% | 13.37% | 0.24% | 1.86% | 2.10% | 88.43% |
| Austria | 0.60% | 3.54% | 4.25% | 83.29% | 16.49% | 0.60% | 3.05% | 3.76% | 81.13% |
| Germany | 0.66% | 4.34% | 5.00% | 86.84% | 20.56% | 0.66% | 3.88% | 4.54% | 85.50% |
| Iceland | 0.21% | 0.37% | 0.58% | 63.82% | 10.66% | 0.21% | 0.49% | 0.70% | 70.11% |
| Israel | 0.38% | 1.81% | 2.16% | 83.75% | 19.81% | 0.38% | 1.78% | 2.16% | 82.55% |
| New Zealand | 0.22% | 1.33% | 1.55% | 85.71% | 17.08% | 0.22% | 1.33% | 1.55% | 85.71% |
| South Korea | 0.59% | 2.06% | 2.65% | 77.73% | 18.63% | 0.59% | 2.07% | 2.66% | 77.80% |
| Mean | 0.41% | 2.11% | 2.54% | 80.82% | 16.66% | 0.41% | 2.07% | 2.50% | 81.60% |
| SD | 0.20% | 1.37% | 1.57% | 8.04% | 3.56% | 0.20% | 1.11% | 1.30% | 6.14% |
| 95% CI of Mean |  |  |  |  | (12.92%, 20.4%) |  |  |  |  |

### Calculation of the variation in each country’s corrected CFR values due to the case population age distribution

We next tested how much of the variation in the corrected CFR between countries was due to their age distribution. The corrected CFR was broken into two components, one from cases 70 years old and above (cCFR_70+_) and the other from cases 69 years old and below (CFR_0–69_).

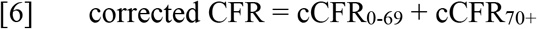

The terms in Eq. [6] were calculated for each country from the following expressions:

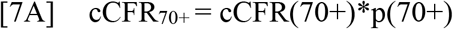

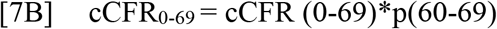

To determine how much of the variation in cCFR_70+_ between countries that can be explained by p(70+), we calculated the R^2^ of the least squares regression.

We further broke down cCFR_70+_ to understand how much of the variation could be explained by the population in the 70–79 age group and 80+ age groups separately using Eq. [7C]:

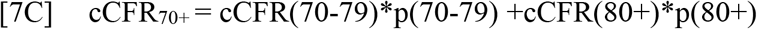

We adjusted for the percentage of cases who were 80+ out of the percentage of cases age 70+, 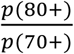, as shown in Eq. [8A]. The reported values for 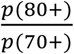 ranged from 0.24 to 0.53 with a mean of 0.40 (Table 3). In order to assess whether the remaining variation in cCFR_70+_ was due to this range in 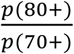 we calculated an adjusted value of cCFR_70+_ for each country by normalizing 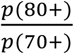 to 0.40 as given in Eq. [8B].

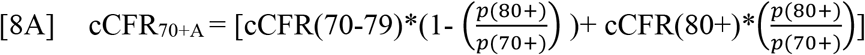

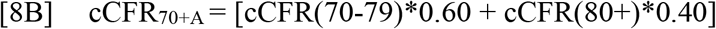

To determine how much of the variation in cCFR_70+A_ between countries can be explained by 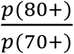, we calculated the R^2^ of the least squares regression.

### Simulation of the closed case CFR(t)

To understand the basis for the apparent early convergence of the closed case nCFR to the corrected CFR value, we calculated the cumulative number of recoveries versus day after the outbreak using the above approach for calculating cumulative fatalities (Appendix 5). Case per day data from South Korea and Germany were used in the simulations. Based on recent reports (Verity, Bi) and earlier work by Ghani with SARS, the distribution function for time to recovery *f*_R_ is similar to that for fatality but with a median shifted several days later and a less right skewed distribution.(1,5,32) Based on these reports, we used a lognormal *f_R_* with a logSD of 0.25 and examined the effect of the median shift on the convergence to the corrected CFR value of closed case CFR(t) curves.(5,32) The closed case CFR(t) was calculated using the following formula,

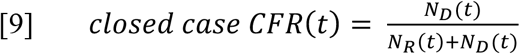

### Calculation of the percentage of the adult population of New York City that has been infected with COVID-19 on April 22, 2020

We used our model to estimate the true corrected CFR for New York City using the reported percentages of cases above 0–69, 70 – 79, and 80+ years old in Eq. [5] with the CFR() coefficients derived from the countries we analyzed.(31) We then estimated the true IFR by dividing by 2, based on reports that up to half of the infected cases in Iceland and the Diamond Princess cruise ship are asymptomatic.(6,13,34)

To estimate the total number of infected individuals in the population, we divided the time corrected number of fatalities by the IFR.(31) The time correction factor (CF_t_) of 1.74 was calculated from the new cases per day as described above. We assumed based upon a relatively constant number of tests per day over this period that the captured cases would be proportional to the total number of new cases per day in the population.(31,36,37)

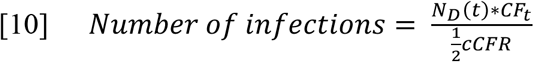

For the total number of fatalities, we used the confirmed cases to attain a minimum estimate; we then added probable fatalities for a maximum estimate. To determine the percent of the adult population infected, we then divided the maximum and minimum number of infections by the number of adults (over age 18) in New York City.(36) The adult population number was used due to the random testing not including children, who are known to have a much lower symptomatic and total infection rate than adults.(13,24–27) We also compared our calculations with other models using their reported IFR values (Table 1) and Eq. [10].

## Results

### Increase in the reported nCFR(t) versus day after the start of the outbreak in five countries

We found in all countries examined that the reported nCFR has risen throughout the COVID-19 outbreak. As shown in Fig. 1, the value of the reported nCFR(t) for Germany rose from a low value of 0.12% on March 10, 2020 to its present value of 4.36% on May 7, 2020. Our estimate of the final CFR of 5.0% is shown as a dashed horizontal line. Figure 2 shows a similar plot using the South Korean data, which rose from a low of 0.55% on March 8, 2020 to its value on May 7, 2020 of 2.28%. The values shown are plotted from 10 days after the first 100 cases were reported to avoid large fluctuations, these due to the low number of fatalities early in the pandemic. In Appendix 3, we show that the nCFR versus day curves for Austria, Australia, Iceland, Israel, and New Zealand exhibit the same trend.

**Figure 1.**
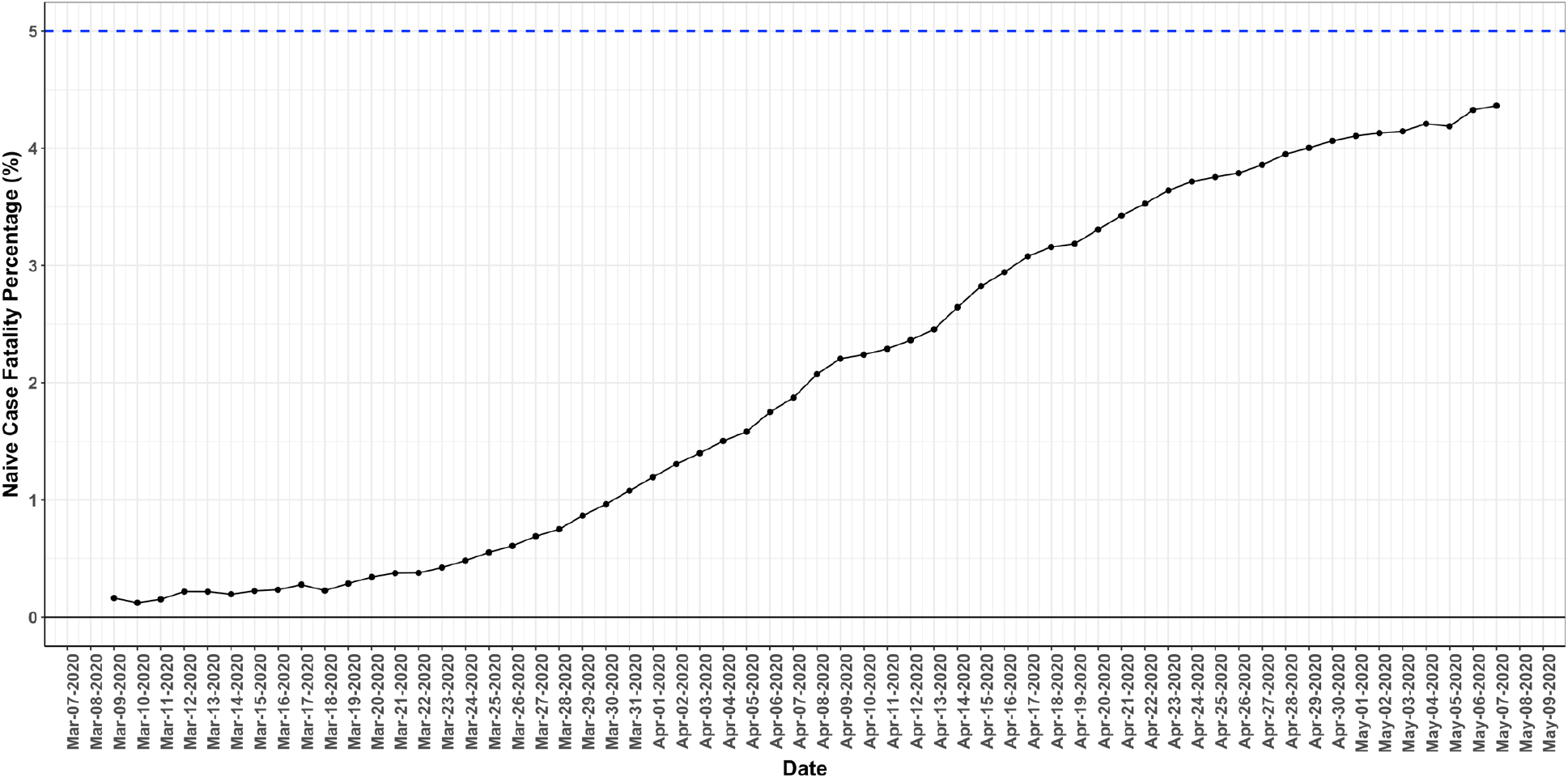
Reported nCFR(t) increases with time after outbreak for Germany. The nCFR has continuously increased with time after the outbreak, from a lowest value of 0.12% on March 10, 2020 to its present value on May 7, 2020 of 4.36%. The dashed horizontal line at 5.0% is our estimate of the final CFR from the closed case CFR value.

**Figure 2.**
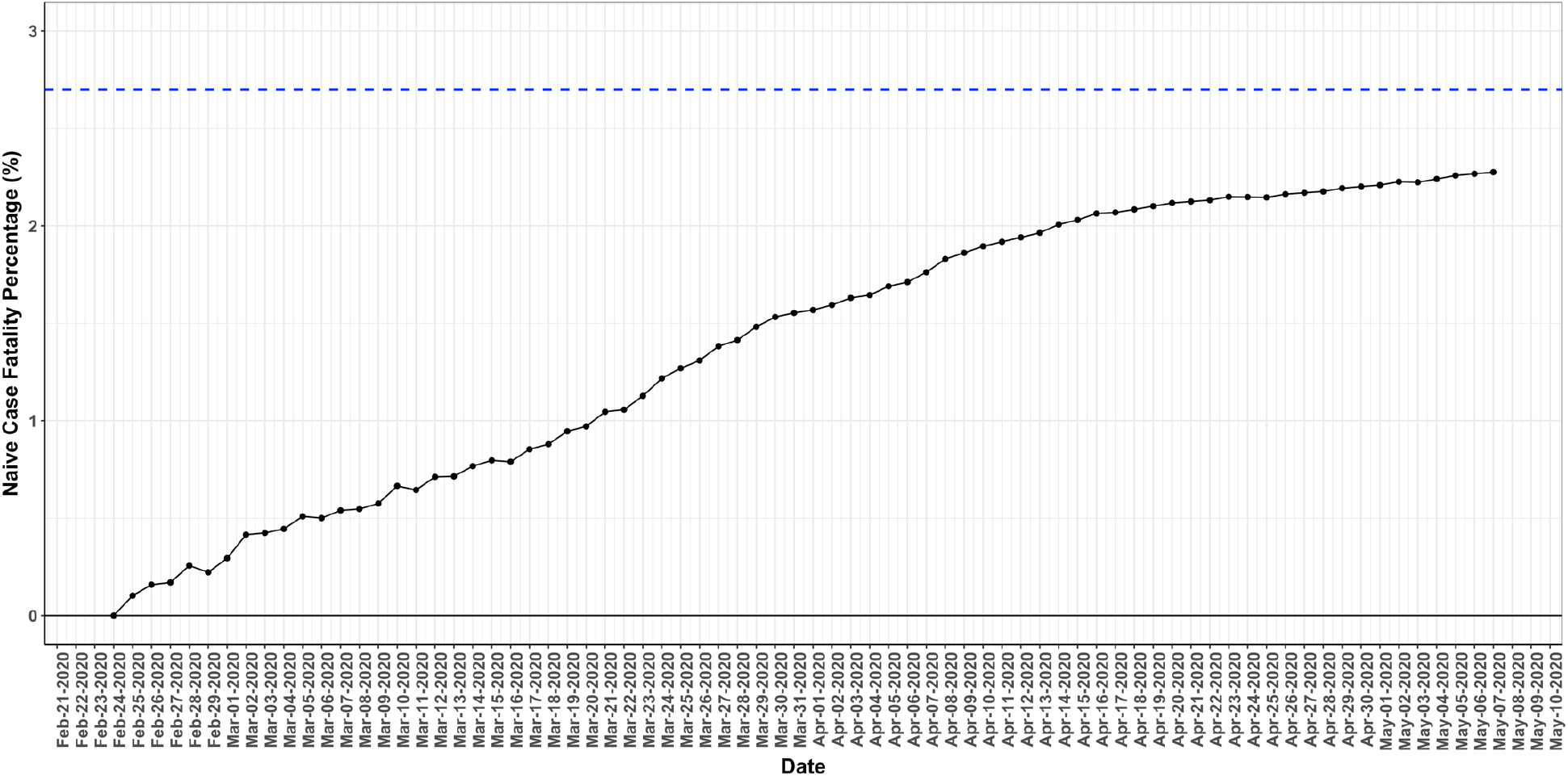
Reported nCFR(t) versus day after outbreak for South Korea. The nCFR for South Korea is also seen to be continuously increasing from a minimum of 0.55% on March 8, 2020 to 2.28% on May 7, 2020. Our estimate of the corrected CFR of 2.7% is shown as a dashed blue horizontal line.

### Increase in number of fatalities on the Diamond Princess cruise ship since February 17, 2020

The Diamond Princess cruise ship is unique in that a large contained group had a significant number of infections and were all tested. Early reported nCFR values of this and other cruise ships with outbreaks have been used to support low corrected CFR values.(4,8) Russell et al. (2020) analyzed the number of fatalities per day from Feb. 17, 2020 through March 3, 2020 and using a similar time correction approach to ours, we estimated that the final nCFR (equivalent to corrected CFR since all cases were tested) would be 2.1% with 15 fatalities.(6) Figure 3 plots the cumulative number of fatalities on each day after the ship docked. As of April 17, 2020, the number of fatalities is now 15 in agreement with this prediction.(6)

**Figure 3.**
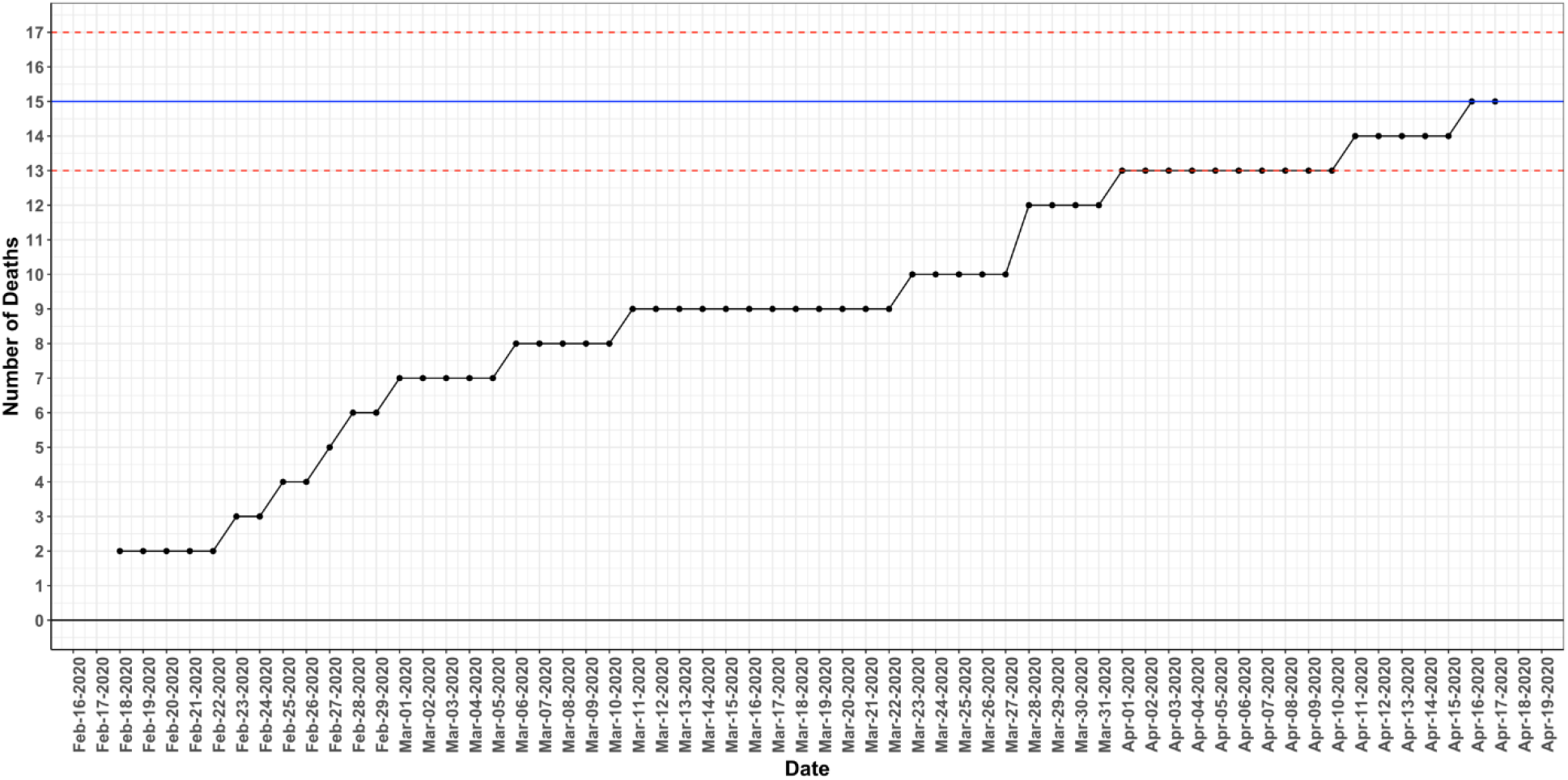
Total number of fatalities from February 17, 2020 to April 17, 2020 for the Diamond Princess cruise ship. The solid horizontal line is the number of fatalities predicted if the true CFR was 2.1% as predicted by Russell et al. using a similar time to fatality correction as (to) ours.(6) The red dashed lines are the 95% CI for the estimate. As with the seven countries examined, the number of fatalities continues to increase with time.

### The reported closed case nCFR converges to a constant value before the nCFR

We found that for the countries we examined, the closed case CFR appears to have converged to a constant value prior to nCFR. In Figure 4, we plot the reported closed case nCFR(t) and nCFR(t) curves from Germany as an example, showing clearly that the closed case nCFR has converged 48 days prior to May 7, 2020, while the nCFR continues to increase. Appendix 2 shows that a similar convergence has occurred for Australia, Austria, Iceland, Israel, New Zealand, and South Korea.

**Figure 4.**
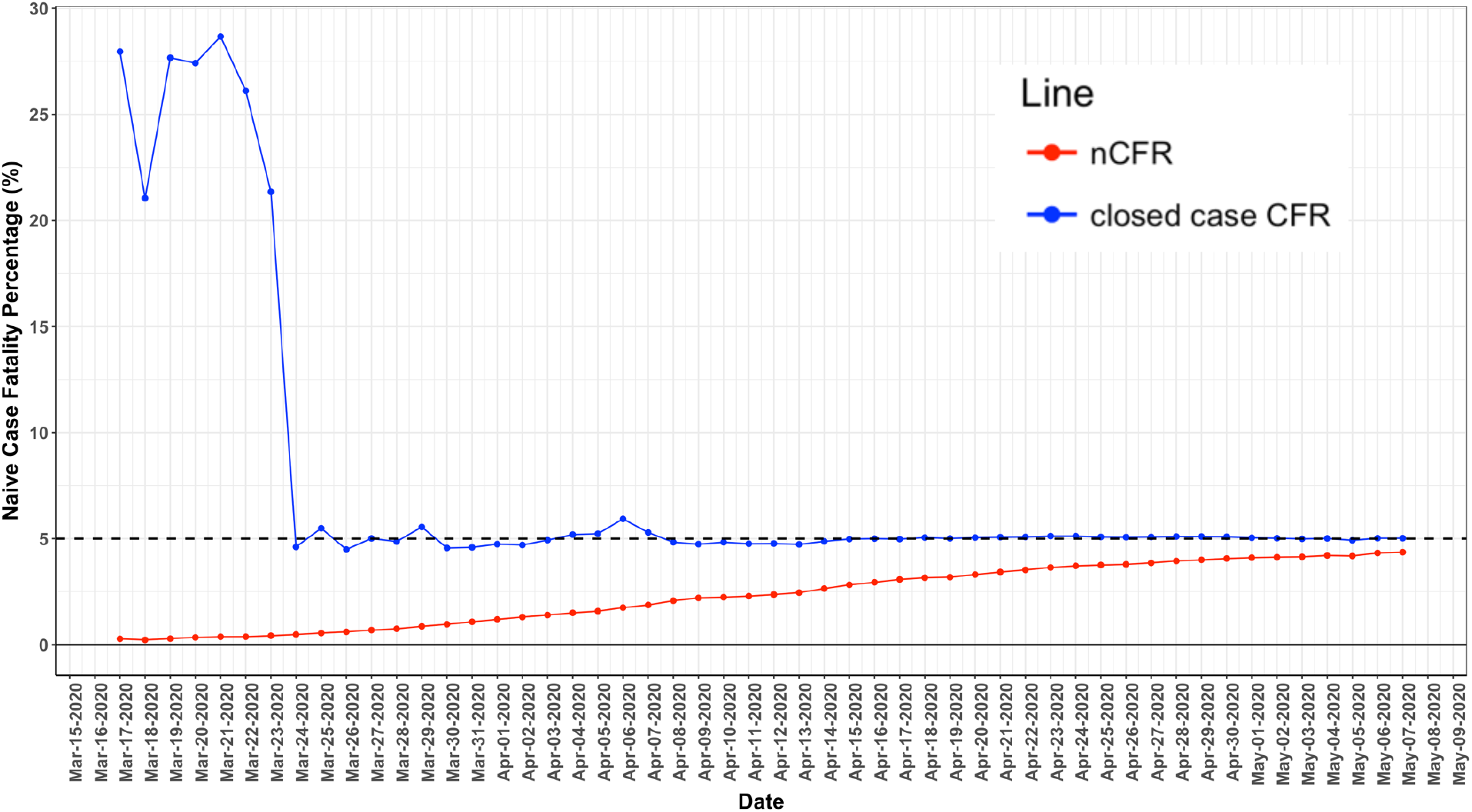
Closed case CFR(t) and nCFR(t) versus day for Germany. As shown by the top curve, the closed case fatality ratio appears to be converging to a near constant value. In contrast the nCFR continues to climb, and as of May 7, 2020, is still well below the closed case CFR.

### Estimation of the corrected CFR from the closed case CFR and time to fatality correction of the reported nCFR

As described in Calculations, we calculated the corrected CFR using two methods. As shown in Table 2, the closed case CFR and standard time correction methods gave similar results for all of the countries examined.

### Assessment of the accuracy of determining closed case CFR by time correction early in an outbreak when the reported nCFR is several fold below the final value

In order to determine whether the nCFR(t) curve could be used throughout to predict an accurate corrected CFR value, we performed simulations in which we generated predicted nCFR(t) curves for different CFR values. Figure 5A and 5B show the simulated nCFR(t) and N_D_(t) curves for Germany using corrected CFR values of 0.5%, 1.0%, 2.0%, 3.0%, 4.0%, 5.0% (the final corrected CFR value denoted by an asterisk), and 6.0%. It is seen that the simulated nCFR(t) curve using this value is in sound agreement with the reported nCFR(t) data, even when the reported nCFR was well below 1.0%. In contrast, the curves consistent with previous corrected CFR estimates in the 0.5% to 1.0% range (Table 1) poorly fit the reported nCFR(t) data at all times. We found similar results for the other countries analyzed (Appendix 2), although with more variation in countries with low numbers of total cases (e.g. Australia, Iceland, New Zealand), possibly due to statistical fluctuations in the number of fatalities at early times.

**Figure 5A.**
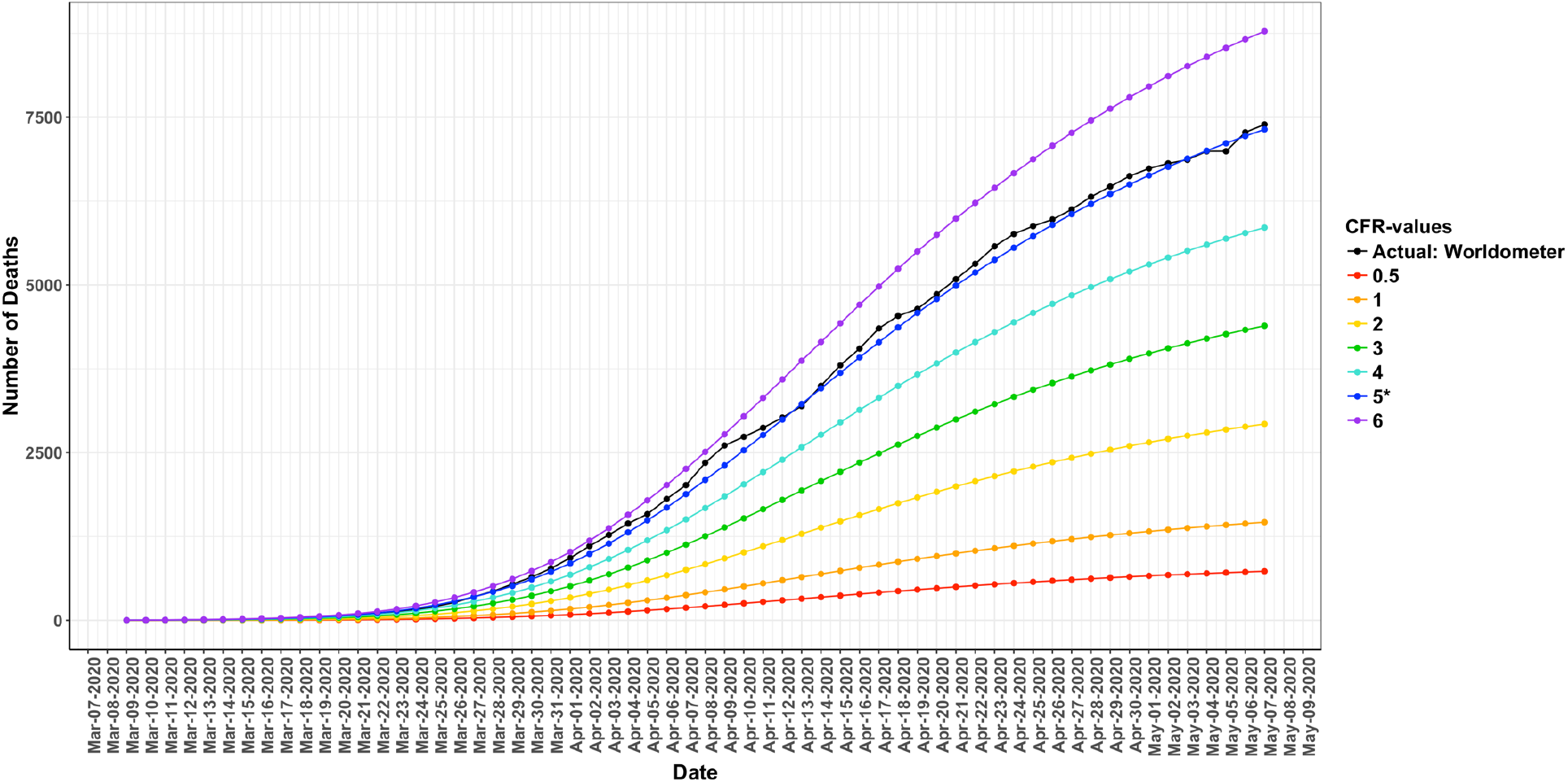
Simulated and reported N_D_(t) versus day curves for the Germany. The black curve is the reported data. The best match to the reported data throughout most of the outbreak is for the final corrected CFR of approximately 5.0%, which is the same as the measured closed case CFR on May 7, 2020.

**Figure 5B.**
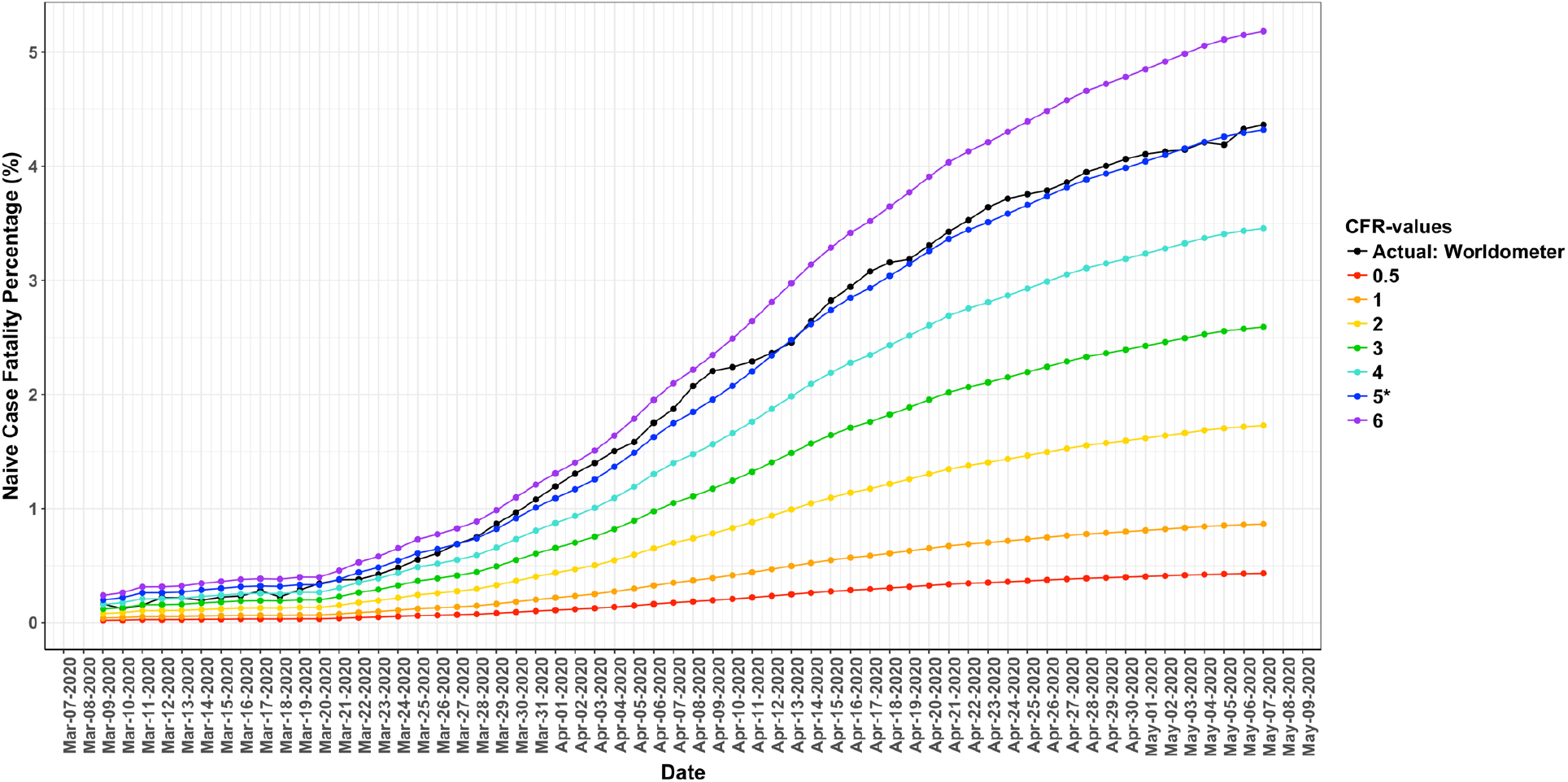
Simulated and reported nCFR(t) versus day curves for Germany. The reported nCFR(t) curve is plotted in black. Even though the reported nCFR(t) curve rises continuously, it is well matched throughout the duration by the calculated nCFR(t) curve (blue), with the final corrected CFR of 5.0%. This corrected CFR is over 10-fold higher than the reported nCFR values early in the outbreak.

### Determination of how much of the variation in the corrected CFR between countries can be explained by the percentage of cases 70 years old and above

Several reports have indicated that there is a sharp increase in the CFR from COVID-19 above approximately 70 years old and these cases account for the majority of fatalities.(6,15) However, the reported values may be biased due to countries prioritizing the testing of high risk individuals in older age ranges. In order to assess these values under the conditions of minimum bias in testing we tabulated (Table 3) for each country the values of the corrected CFR for each age group (CFR(0–69), CFR(70- 79), CFR(80+), CFR(70+)), the percentage of the case population in each group (p(0–69), p(70- 79), p(80+), p(70+)) and the contribution of each group to the overall corrected CFR (CFR_0–69_, CFR_70–79_, CFR_80+_, CFR_70+_).

Figure 6A plots the two components of the cCFR for each country comprising cases 0–69 years old (cCFR_0_-_69_, green) and the cases 70 years old and above (cCFR_70+_, blue) versus p(70^+^). It is seen that for all countries, cCFR_70+_ is the major component of the CFR with the ratio cCFR_70+_/cCFR having a mean value of 81% with an SD of +/− 8% (Table 3, note that this is also the ratio of total fatalities in the 70+ age range to total fatalities). To determine how much of the variation in this component could be explained by the percentage of cases 70 and above, as opposed to variations in testing, we performed a regression analysis against p(70+) giving:

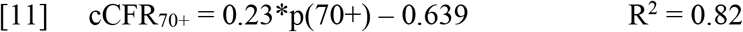

**Figure 6.**
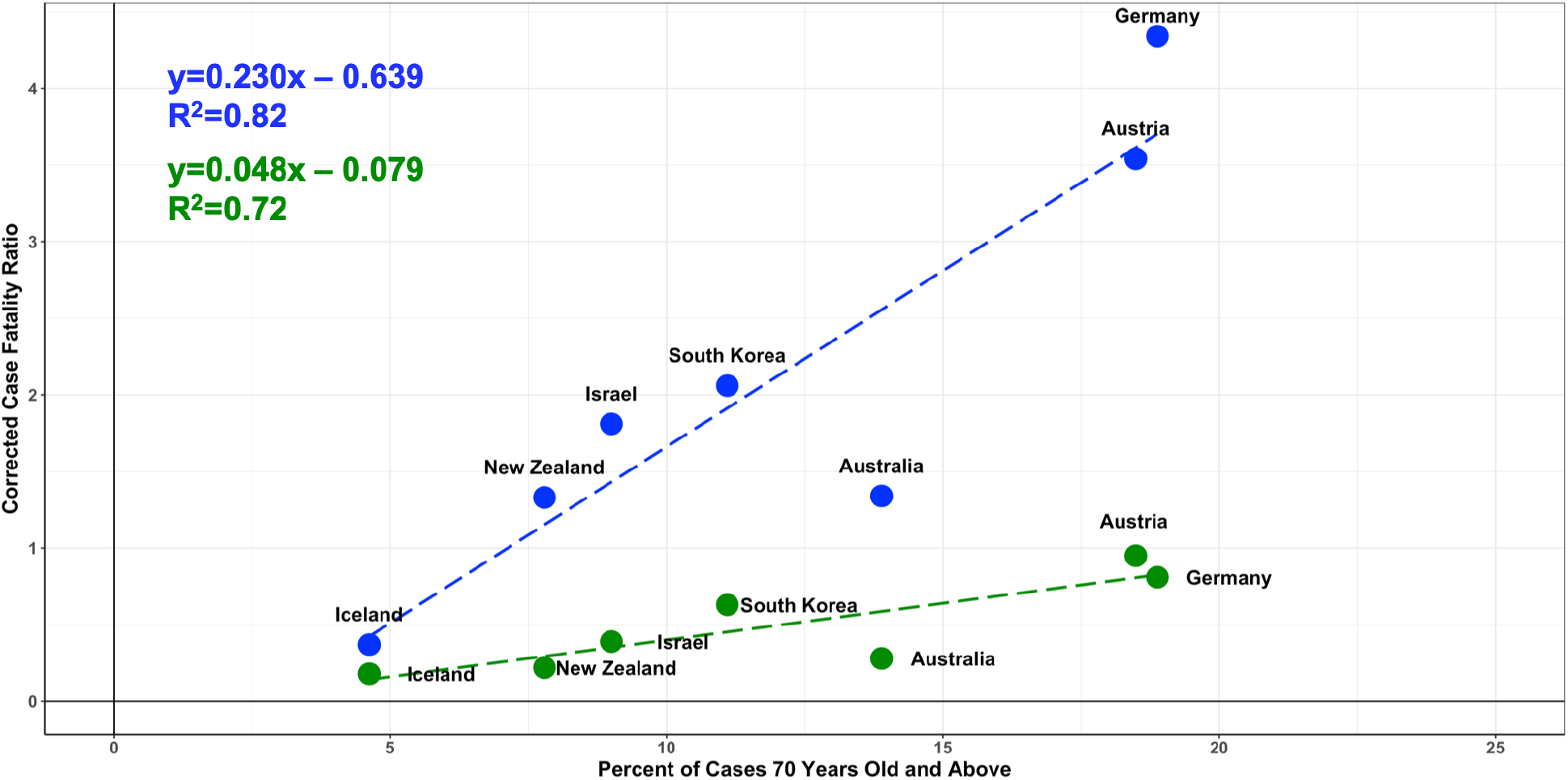
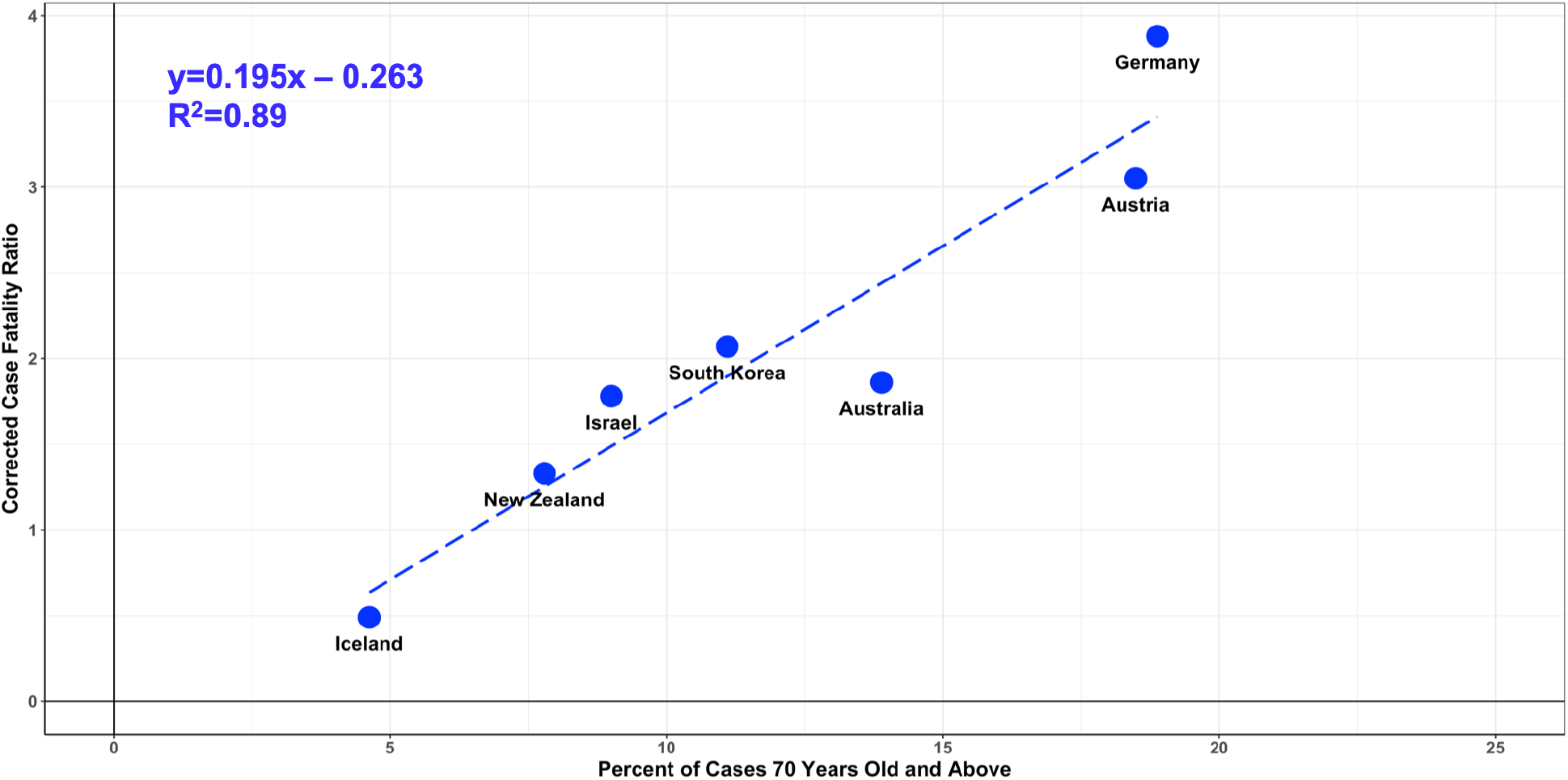
Linear regression analysis of cCFR_70+_, cCFR_70+A_, and cCFR_0_–_69_ versus percent of cases 70 years old and above (p(70+)). **Figure 6A:** shows a plot of cCFR_70+_ (blue) and cCFR_0_-_69_ (green) for each country versus the percent of cases 70 years old and above (p(70+) The total corrected CFR for each country is given by cCFR = cCFR_70+_ + cCFR_69_. It is seen that for all countries the cCFR_70_ term explains the large majority of cCFR (81% +/− 8%). The majority of the variance in cCFR_70_ is explained by cases 70 years old and above (R^2^ = 0.82). The contribution to cCFR from cases 69 years old and younger showed a weak dependence on p(70+) (slope = 0.05, R^2^ = 0.72). **Figure 6B.** shows a plot of cCFR_70+A_ (blue) for each country. The value of cCFR_70+A_ for each country was calculated by adjusting the fraction of cases in the 70 and over group who are 80 years old and above to be 40% (p(80+)/p(70+) = 0.40), which is the mean of the countries examined (Table 3). The higher fraction of the variance explained by age for cCFR_70+A_ (R^2^ = 0.89) indicates that cases 80 years and over are an important factor in the overall cCFR.

In order to assess whether additional variation could be explained by the relative percentage of cases 80 years old and above (p(80+)) in the 70+ age group, we calculated using the measured cCFR(80) values and adjusted cCFR_70+_ for each country (cCFR_70+A_) for a constant ratio of p(80+)/p(70+) of 0.40. This ratio was chosen to be the mean value of the seven countries (Table 3). As shown in Figure 6B, linear regression of cCFR_70+A_ against p(70+) showed a reduction in variation relative to Eq. [11].

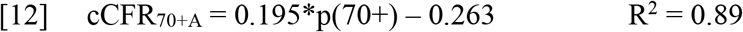

In order to further test the importance of age in explaining the variation between countries we calculated the slope for Eq. [12] using the values of CFR(70–79) and CFR(80+) from each country and Eq. [8]. This method gave a mean slope of 0.167 with a 95% CI of the mean of 0.129– 0.204, which is within error the same as the slope determined by linear regression value.

Although the age group below 70 years old only accounted for on average 19.1% of the fatalities (and the same percentage of the total cCFR as shown in Fig. 6A), we still found that this group was a significant factor in overall mortality, with an average cCFR_0–69_ being 0.41% (Table 3). There was also weak dependence of cCFR_0–69_ with p(70+) (slope = 0.05, R^2^ = 0.72).

### Estimation of the corrected CFR for China as of February 11, 2020 and New York City as of April 22, 2020

We applied the linear model described in Eq. [5] to predict the corrected CFR based on the age distribution of cases in China on February 11, 2020 (WHO).(2) Based on the percentage of cases from 70 to 79 (9%) and 80 and above (3%), the model predicted a corrected CFR of 2.19% with a 95% CI of the mean: 1.54%-2.85%. We performed a similar calculation based on the age distribution of positive cases in New York City as of April 22, 2020 with 9% of cases between 70 and 79 years old and 8% 80 years old and older.(31,36,37) Due to the greater percentage of cases in both categories, the corrected CFR for New York City was predicted to be 3.60%, with a 95% CI of the mean: 2.73%-4.47%.

### Estimation of the percentage of the population infected with COVID-19 in New York City

As described in the Calculations, in order to validate the model, we calculated the percentage of the New York City population that has been infected by COVID-19 up through April 22, 2020 in order to compare with recent studies that have performed random testing. We calculated a maximum and minimum value based on whether unconfirmed but probable fatalities were included. The inset shows our minimum and maximum calculated values (green bars) of 14.69% (95% CI of mean: 11.85%–19.43%) and 22.05% (95% CI of mean: 17.75%– 29.10%) are in agreement with two recent studies that randomly tested individuals in the NYC adult population of 15.3% and 21%, respectively (blue bars).(15,16) Also plotted are the predictions from previously reported true IFR calculations that have been applied to estimate fatalities in the United States and United Kingdom (Table 1). For the other IFR values only confirmed COVID- 19 fatalities were included. As seen in the graph, other than Modi (2020) and the Imperial College model, the estimated percentages of the population infected are several fold above the reported values.(5,11)

## Discussion

To obtain an estimate of the true CFR for COVID-19, we focused on countries with extensive testing of symptomatic and asymptomatic individuals (Appendix 4), and with access to modern medical care. We assumed that the corrected CFR from these countries represents a conservative estimate of their true CFR, without the need to introduce potentially large corrections for missed cases.(3–11,20–22,34) We found that in all cases there was a several fold increase in the reported n CFR, which could be accounted for by the same distribution function describing the delay between diagnosis and fatality. Despite the high level of testing we found a wide range of corrected CFR values for these countries (from 0.7% to 5.0%). However, on average 81% (95% CI of mean: 72.39%– 89.26%) of the total CFR was due to the cases 70 years old and above. When we examined the component of the CFR due to this population (cCFR_70+_) separately, we found that 89% of its variance was explained by a linear model based upon the percentage of cases 70–79 and 80 and above years old. The remaining component of the corrected CFR was relatively stable between countries with a mean value of 0.41% and 95% CI of the mean of 0.21%-0.62%. We tested the model by using it to calculate the true CFR and IFR for New York City where there is a large infected population that has recently undergone several studies performing random testing. Our estimated corrected IFR value for New York City of 1.80% as of April 22, 2020 was 2 to 16.5-fold higher than previous values that have been applied to the US and UK (Table 1). However, its prediction that between 14.69% and 22.05% of the New York City adult population has been exposed to COVID-19 gave the best agreement with the results of random testing studies in New York City at or near April 22, 2020 (15.3% – 21%, Figure 7).(15– 17) In contrast, previous IFR values used in epidemiological models for forecasting fatalities in the United States and United Kingdom predicted values between 1.5 and 10 fold higher. We discuss below the potential sources of difference between our study and previous CFR and IFR calculations, and the relative strengths and limitations of our approach.

Our calculated corrected CFR for China was 2.19% with a 95% CI of the mean: 1.54%-2.85%, which was considerably lower than for New York City due to the lower fraction of cases between 70 and 79 and 80 and over years old, but higher than the majority but not all of the previously reported values (Table 1), which ranged from 0.9 to 5.6%. Almost all of the studies that calculated an IFR/CFR using data from China performed a time correction similar to that applied here, so that it is unlikely that the time delay to fatality significantly contributed to their lower corrected CFR values.(3,5–7,20–23) A more likely factor explaining the differences between values is the correction for missed cases, which ranged from 1.0 to approximately 4.(20,23) In addition to uncertainties in determining missed cases, a significant amount of the difference can be accounted for when previous values are scaled up by 1.5 fold to match the recent report of the Chinese government of undercounted fatalities.(2)

The reported nCFR(t) versus day data shows clearly that the nCFR(t) values have all risen at least several fold from the early values used to justify low estimates of the IFR for COVID-19 (see Figures 5A, 5B, and Appendix 3).(12,14,24–27,30) We therefore calculated the corrected CFR for each country using both the earlier convergence of the closed case CFR to a final value, and a conventional time to fatality correction method.(5–7,23,32,33). In all cases the corrected CFR calculated using both methods were similar (Table 2). As shown in Figure 2, the closed case CFR for Germany converged to its final value 44 days before May 7, 2020, while the reported nCFR is still rising, and similar early convergence was seen in all of the other countries evaluated (Appendix 1). The simulations in Appendix 5 show that the convergence time to the final corrected CFR value for the closed case CFR primarily depends on the separation of the median days to fatality versus the median days to recovery with the fastest convergence when the medians are the same. We also found that the time to fatality correction method for calculating the corrected CFR worked well at early times when the reported nCFR values were several times higher than the reported values on May 7, 2020. Therefore, both methods should be useful early in an outbreak to provide a more accurate measure than the uncorrected nCFR.

We did not factor in preexisting conditions in our analysis which has been reported as significantly affecting mortality.(15,17,26,31,36,37) The high R^2^ values we found for the cCFR_70+_ component (0.82 and 0.89) suggests that other risk factors than age were similar between the countries we analyzed. However, it is likely that the ability to extrapolate from our findings to other countries would be improved if preexisting conditions and other risk factors that may be differentially present were included in our model.

The corrected CFR between countries for the under 70 years old component (cCFR_0_-_69_), was considerably lower than the cCFR_70_ in all cases, mean 0.41% with a 95% CI of the mean: 0.21%- 0.62%. It is likely based on other reports that the variation in cCFR_0–69_ could be explained by the fraction of the case population in the 60 – 69 group which also has a highly elevated risk.(13,24–27) However due to the low number of fatalities in several countries in the 0– 69 age groups we did not perform a sub analysis.

To calculate the IFR for New York City, we divided the calculated corrected CFR by a factor of 2, based on reports from the Diamond Princess and Iceland, that half of COVID-19 cases are asymptomatic. This value may be an overestimate, as shown by Mizumoto, because these reports did not take into account the lag between infection and the onset of symptoms, which led him to revise the true asymptomatic case number for the Diamond Princess to approximately 30%.(34) A potential confound in our approach is that the time correction of the reported number of fatalities is based upon the assumption that the percentage of missed cases was constant, since all symptomatic cases were not captured. The validity of this assumption was supported by data from the New York City Department of Health that the number of tests per day was close to constant during the period up to April 22, 2020.(31,36,37) We note that in the extreme alternate possibility in which none of the active cases as of April 22, 2020 subsequently died the percent of infections calculated would range from 8.6% to 13.0%.

Our calculation of the minimum and maximum percentage of the adult population in New York City that has been infected by COVID-19 agreed with the recent studies that performed random testing of segments of the adult population (Figure 7).(15–17) In one study, 15.3% of women entering two New York City hospitals to give birth were found by testing to be infected with COVID-19 (33 out of 215 having the virus).(16) In the second study the New York City infected population was estimated at 21%, this from 3000 serological antibody-based measurements of passersby at testing stations near public areas in New York City and other regions in New York State (with the results reported on April 22, 2020).(15) The New York City findings were replicated from subsequent testing of 5500 cases reported April 28, 2020 (24% infected) and 15, 500 cases reported on May 2, 2020 (19.9% infected). Due to the heterogeneity in COVID-19 fatalities and cases within even New York City, and due to the restricted age range of the groups examined (18 – 75 for the New York State study), these percent infection values may be overestimates.(31,36) However, given that the large majority of cases in New York City are between ages 18 and 75, it is unlikely that this bias would have a large impact.

**Figure 7.**
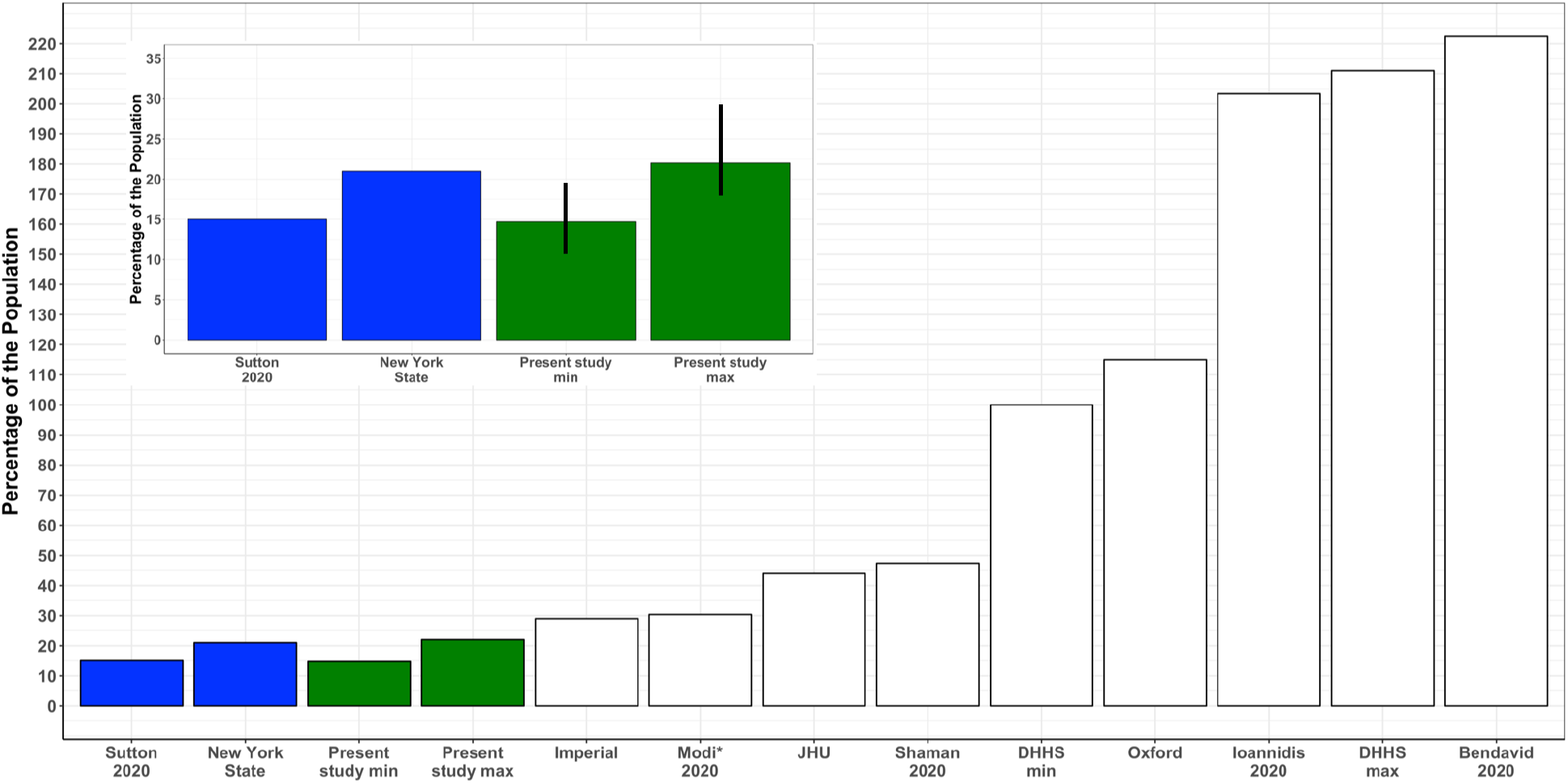
Reported percentage of New York City adults infected with COVID-19 versus calculated using our and other reported IFR values. As shown in the inset, the predicted maximum and minimum percent of the population in New York City infected with COVID-19 is in logical (good) agreement with recent reports based on random testing.(15,16) For comparison, we calculated the percentage of the population infected, and then predicted using previous reported IFR values for New York City, the United States, and the United Kingdom (see Table 1).

As shown in Figure 7, the predicted infection percentage for the adult New York City population using the IFR values from previous models was between 1.5 and 10-fold higher than the actual reports as well as our predictions. The closest IFR values to ours were reported by Modi et al. and the Imperial College model.(11,19) The model of Modi estimated the New York City IRF based upon the difference between reported fatalities during the outbreak versus a similar period of time, and corrected for age distribution of positive cases.(11) No time correction was used to correct the IFR due to the investigators wanting to report a most conservative value. To compare with our predictions, we applied the same time to fatality correction to the Modi IFR that we used in our model. This increased its IFR to 0.88%, similar to the value of 0.90% used in the Imperial College model.(5)

Much lower estimates of the IFR have recently been reported based on serological antibody testing in Santa Clara County in early April of 0.13%–0.2%.(8) A potential explanation for the difference from our estimates is that the actual percentage measuring positive in their study was low, between 2.49% and 4.16%. A recent study that has evaluated available serological testing platforms found that the fraction of false positives can exceed this range.(38) The impact of false positives is likely to be less significant for the New York State study because of the much higher true percentage of the New York City population that is infected. Additional validation of the New York results is from their finding consistently of low infection percentages (∼ 1.0%) in several regions in New York State outside of the New York City metro area which supports a relatively low false positive rate.(15,17,31)

Having an accurate value of the true CFR and IFR is vitally important for modeling the total number of fatalities from COVID-19. As shown in Figure 7 the IFR values used in several leading epidemiological models are not compatible with the number of infections in New York City. It has been shown that an increase of IFR from 0.25% to 0.5%, approximately doubled projected fatalities with other factors held constant.(28) Although our approach is less sophisticated than missed case correction it explains the majority of the variation in the countries examined and is consistent with the most reliable random population testing study available. Therefore, it should be valuable for providing at conservative upper CFR and IFR bound for epidemiological models of the COVID-19 pandemic.

## Data Availability

The data is available upon reasonable request to the authors.

## Acknowledgments

The authors acknowledge invaluable assistance from Julia Rothman in harvesting the time course data used to perform the analysis from multiple sources. Gerard Bossard provided expert review and editing of portions of the manuscript. DLR acknowledges helpful suggestions for the paper from Gail Rothman, John Rothman, Jeff Evelhoch, Gerard Sanacora, Kevin Behar, Marcia Johnson, Barbara Gulanski and Anthony Basile. The authors did not receive support for this work.

## Notes

### Competing Interest Statement

The authors have declared no competing interest.

### Funding Statement

The authors did not receive any external funding for this work.

### Author Declarations

All relevant ethical guidelines have been followed.

